# Magnitude Of Birth Outcome After Induced Labor Among Mothers Delivered In Nigist Eleni Mohammed Memorial Comprehensive Specialized Hospital, Hosanna Town, Hadiya Zone, Southern Ethiopia. Cross-Sectional Study

**DOI:** 10.1101/2023.07.20.23292943

**Authors:** Dereje Worku, Tesfaye Assebe, Tesfaye Gobena, Temesgen Kechine

## Abstract

**Background:** “Induction of labor is not risk free, despite its importance for ending risky pregnancy compare to spontaneous onset of labor it has potential harms and it increases the rate of different maternal and neonatal complications.” Due to this WHO recommends IOL with only clear medical indications when the benefit more significant than potential harms. Even though there is a few study on IOL that address magnitude of birth outcome after IOL no study is found that determine the contributing factors to birth outcomes after IOL in Ethiopia especially in my study area, therefore the factors that affect the birth outcome after IOL need to be clearly understood.

**Objective:** To assess the magnitude birth outcome after induced labor and associated factor among child bearing mother who deliver in NEMMCSH in the last two years (January 01, 2019 to December 31, 2020 GC). Data was collected from June 25 to July 09, 2021 GC.

**Methods:** Hospital based retrospective cross-sectional study was conducted on 778 study participants selected by systematic random sampling technique among all child bearing mothers delivered by induction in NEMMCSH from January 01, 2019 to December 31, 2020 GC. Data was collected from patient cards, delivery registration log books and operation note books. Then data were entered and coded using EPI data version 3.1 and analyzed using SPSS version 25. Bivariate and multivariate logistic regression analysis was carried out to determine the association different potential factors with the birth outcome after IOL. Independent predictors were determined using adjusted odd ratio with 95% CL at p value < 0.05 in multivariate logistic regression analysis.

**Results:** In this study the magnitude of still birth after IOL was 9.6%. Rural residence [AOR=3.59; 95%CI:(1.32, 9.80)], maternal chronic medical diseases [AOR=3.58; 95% CL: (1.23, 10.41)], history of previous still birth [AOR=7.45; 95%CI: (2.45, 22.38)], Partograph use [AOR=0.034; 95%CI: 0.01,0.09)], delivering ˂8 hours[AOR=0.13; 95%CI: (0.03,0.56)] and delivering within 8-16[AOR=0.28; 95%CI:(0.10, 0.76)] hours were significant predictors for still birth.

**Conclusions:** The magnitude of still birth after IOL was relatively high in the study area. Variables which increase the likelihood of still birth were, living in rural area and previous history of still birth. The recommendations also forwarded for health care provider, NEMMCSH, different stakeholders and for researchers.

## Introduction

“Induction of labor is defined as the process of artificially stimulating the uterus to start labor. It is usually performed by administering oxytocin or other prostaglandin to the pregnant women, or by artificially rupturing of the amniotic membrane” (67)

During compilations of pregnancy (PROM, pre-eclampsia, IUGR, and post term pregnancy) that confer significant ongoing risk for the fetus and the mother IOL is often the most important medical intervention utilized to decrease the neonatal and maternal morbidity and mortality (18).

IOL can be planned (elective) or emergency. Elective induction is usually done with prior planning by health care provider and the mother when continuing the pregnancy beyond certain weeks has risk for the mother or the fetus, in such case of, PROM, DM, hypertension, postdate pregnancy, small or large for baby. Emergency induction is done when there is an emergency maternal or fetal conditions that necessities induction of labor immediately in case of pronged PROM, severe IUGR, and intrauterine infection, pregnancy beyond 42 weeks and eclampsia and pre-eclampsia (54). So IOL is not risk free, despite its importance for ending risky pregnancy compare to spontaneous onset of labor it has potential harms and it increases the rate of different maternal and neonatal complications. Due to this WHO recommends induction of labor with only clear medical indications when the benefit more significant than potential harms (65).

A secondary analysis of WHO global survey shows IOL is associated with poor birth outcome such as still birth, low Apgar score at birth and at 5 minutes, low birth weight, need of NICU admission, in Asia and Africa (61). WHO and study in India and Nigeria also support the thought that IOL is associated with poor neonatal and maternal outcomes compared with spontaneous onset of labor (32, 55, and 66). Study in Ethiopia also shows that still birth, low Apgar score at one and five minutes after birth and need of admission to NICU are the common adverse birth outcome after IOL(15). In Ethiopia even if there are limited studies on IOL that address magnitude of still birth after IOL but there is no study that determine predictors of still birth after IOL. So this study can fill the gap by identifying the significant predictors after induction of labor.

A policy of planned induction was found to lower the risk of perinatal death (still birth and neonatal death) by about two-thirds compared to a policy of expectant management. The overall strength of evidence for offering IOL for women over 41 weeks of gestation is in order to lower the risk of CS (34). But Health care provider use appropriate or inappropriate indications for IOL which has an impact on resource for performing induction and on the overall CS rate and perinatal death. Rochelle et al. reviewed the records of 4541 induced pregnancies and found that 15% of induction were either not clinically indicated or not documented (41).

In the study area, induction of labor is a commonly done procedure but there is a limitation in undertaking a study on factors associated with still birth after labor induction in SNNPR region particularly in the study area. Therefore this study will help to fill the gap by identifying demographic factors, obstetrics factors, induction related factors, maternal related factors and labor and delivery related factors. Therefore, the aim of the study is to answer the following research questions :I) what is the magnitude of birth outcome after IOL, II) what are the factors affecting birth outcome after induction in the study area; identify predictors to make recommendations for practice, which we may hopefully reduce complication related to induction of labor.

## Methods

### Study context

The study was conducted on Nigst Eleni Mohammed memorial Comprehensive specialized hospital which is the only specialized hospital in Hadiya zone found in Hosanna town. Hosanna town is found 232 Km away from Addis Ababa, the capital city of Ethiopia in southwest of Ethiopia and 194 Km away from Hawassa, capital city of SNNPR. Nigst Eleni Mohammed memorial Comprehensive specialized hospital is one of the largest hospital which serves not only the zonal populations, but also the neighboring zones and Woredas such as Silte, Gurage, Halaba and Kembata - Tembaro over two million people residing in urban and rural parts of southwest Ethiopia.

### Study design and period

Hospital based retrospective cross-sectional study was employed. The study was conducted from January 01, 2019 to December 31, 2020 GC. The data collection period was from June 25 to July 09, 2021.

### Source population

The source population of the study was all women who undergo IOL after 28 weeks of gestation at Nigist Eleni Mohammed memorial comprehensive specialized hospital from (January 01, 2019 to December 31, 2020 GC).

### Study population

The study population was all systematically selected women undergone induction after 28 weeks of gestation in Nigist Eleni Mohammed memorial comprehensive specialized hospital from (January 01, 2019 to December 31, 2020 GC).

### Eligibility criteria

#### Inclusion criteria

All registered women who had IOL after 28 weeks of gestation in Nigist Eleni Mohammed memorial comprehensive specialized hospital from (January 01, 2019 to December 31, 2020 GC).

#### Exclusion criteria

All registered women who had IOL without full documentation and multiple pregnancy

#### Sample Size Calculation and Sampling Technique

A cross section study was conducted by reviewing records from (January 01, 2019 to December 31, 2020 GC (n=760). Systematic random sampling technique was employed to select the study participants

#### Operational Definitions

##### Still birth

Fetus with no sign of life at birth following induced labor after 28 weeks of gestation but alive before the procedure of labor induction (15).

##### Birth outcome

the life status of the fetus at birth after induction of labor and registered on labor and delivery log book as alive birth or still birth (5, 24)

##### Favorable bishop

the bishop score having the value greater than 6. Unfavorable bishop: bishop score having the value ≤ 6(8, 49).

##### Antenatal booking

mother with even single ANC visit.

##### Maternal medical condition

mothers those who have medical problems such as DM, anemia, chronic hypertensive disorder, cardiac diseases and other chronic disease.

##### Previous history of still birth

mother with at least one previous history of still birth.

##### Partograph use

complete monitoring of the labor progress using Partograph from active phase of labor until delivery of baby.

##### Induction to delivery time

the time it takes the mother from starting induction to delivery of the fetus either vaginally or abdominally (24).

#### Data Collection Tools and Quality Control

The data were collected by secondary data sources (document review) using a structured questioners composed of socio demographic, obstetric, induction related and labor and delivery related variables of women who undergone induction of labor. The questionnaires were prepared in English version from similar study (15, 24, and 40) the data collectors were five BSc midwife and supervised by one MSc midwife to follow the data collection process. The investigators and supervisor were checked data accuracy, consistency and completeness daily.

#### Statistical analysis

The collected data were entered and analyzed using SPSS version 25. The data were cleaned before analysis. Descriptive statistics such as frequency and mean were used to describe the study participants. Crude and adjusted odds ratios (95%) confidence intervals were utilized to assess the determinant factors with the birth outcome after IOL. The criterion for selecting independent variables was set at p-value less than 0.25 in bi variable analysis for final model. To control for potential confounder, a multivariable binary logistic regression was performed. A two tailed p-value < 0.05 was performed to show statistical significant. Model fitness was performed with Hosmer-Lemeshow test. And also multicollinearity was checked using at < 10 variance inflation factor (VIF). There were no variables that have VIF greater than 10.

#### Ethical Considerations

Before conducting this study Haramaya University College of Health and Medical Sciences Institutional Health Research Ethics Review Committee (HU-IHRERC) was secure ethical clearance, and Haramaya University was write Official letter to NEMMCSH. Confidentiality of the data was assured through using anonymous checklist and keeping the data in secured place. In addition the names of the study subjects and treating Health professionals were not included in the checklist. Participation for this study is fully voluntary. Moreover informed voluntary written and signed consent was obtained from the hospital medical director after informing the objectives of the study, the possible risk/benefits, confidentiality and rights to be part of this study.

## Results

### Socio demographic and obstetrics characteristics of the mothers

The mean (±SD) age of the mother in this study was 30.20(±6.279) years. More than half of the mothers 461(60.7%) were in the age group of 20-34years. The majority, 396 (52.1%) of the study subjects were urban dwellers in other way more than half of the mothers 507 (66.7%) were had birth more than one times and 612 (80.5%) of the mothers had ANC follow up at least one times. One hundred thirty six (17.9%), one hundred forty nine (29.6%) and one hundred sixty four (21.6%) of the mothers were had previous history of still birth, previous history of IOL and history of preterm pregnancy. Majority 547(72%) of the new born were had gestational age between 38 to 41 weeks and the mean (±SD) gestational age of the neonate was 38.69(±1.829). More than half of the neonate 454(60.2%) were had birth weight between 2500-3999 grams.

**Table 1:** Socio demographic and Obstetrics characteristics of mothers after induction of labor in NEMMCSH south Ethiopia from January 01/2019 to December 31/2020. (n=760).

| Variables | Categories | Birth Outcome |  | Total |
| --- | --- | --- | --- | --- |
|  |  | Alive birth | Still birth |  |
| Age of the mother | <20 | 52(88.1) | 7(11.9) | 59 |
|  | 20-34 | 423(91.8) | 38(8.2) | 461 |
| | ≥35 | 212(88.3) | 28(11.7) | 240 |
| Residence | Rural | 307(84.3) | 57(15.3) | 364 |
|  | Urban | 380(96.0) | 16(4.0) | 396 |
| Parity | Nulliparous | 234(92.5) | 19(7.5) | 253 |
|  | Multiparous | 453(89.3) | 54(10.7) | 507 |
| Gravidity | Primigravida | 207(92.4) | 17(7.6) | 224 |
|  | Multigravida | 480(89.6) | 56(10.4) | 536 |
| Gestational age before induction | Preterm | 98(89.1) | 12(10.9) | 110 |
|  | Term | 496(90.7) | 51(9.3) | 547 |
|  | Post term | 93(90.3) | 10(9.7) | 103 |
| ANC follow up | Yes | 585(95.6) | 27(4.4) | 612 |
|  | No | 102(68.9) | 46(31.1) | 148 |
| History of still birth | Yes | 88(64.7) | 48(35.3) | 136 |
|  | No | 599(96.0) | 25(4.0) | 624 |
| History of END | Yes | 126(81.8) | 28(18.2) | 154 |
|  | No | 561(92.6) | 45(7.4) | 606 |
| History of preterm pregnancy | Yes | 130(79.3) | 34(20.7) | 164 |
|  | No | 557(93.5) | 39(6.5) | 596 |
| History of previous IOL | Yes | 130(87.2) | 19(12.8) | 149 |
|  | No | 557(91.2) | 54(8.8) | 611 |
| Outcome of previous IOL | Successful | 65(90.3) | 7(9.7) | 72 |
|  | Failed | 62(83.8) | 12(16.2) | 74 |
| History of birth asphyxia | Yes | 44(77.2) | 13(22.8) | 57 |
|  | No | 643(91.5) | 60(8.5) | 703 |
| History of referral | Yes | 120(75.0) | 40(25.0) | 160 |
|  | No | 567(94.5) | 33(5.5) | 600 |

### Induction of labor and delivery characteristics

Of the included 760 study subjects 310(40.8%) undergo induction of labor for the indication of PROM followed by PIH 232(30.5%) while, 12(1.6) pregnancies were either not clinically indicated or not documented and 637(83.8%) of the pregnancies were induced by oxytocin infusion and from 125(16.4) mothers who were induced by misoprostol all are taken virginally. From the total study participants included in this study 2 mothers were developed uterine rupture after induction of labor. In other way from the total 760 induced labor 592(77.9%) were successful. From the total mothers included in the study 100(13.2) of them were had chronic medical disease. Of these 17(17), 11(11), 38(38) and 34(34) were had chronic hypertension, cardiac problem, DM and others medical diseases respectively and the mean (±SD) time for induction to delivery was 13.20±4.943.

**Table 2:**
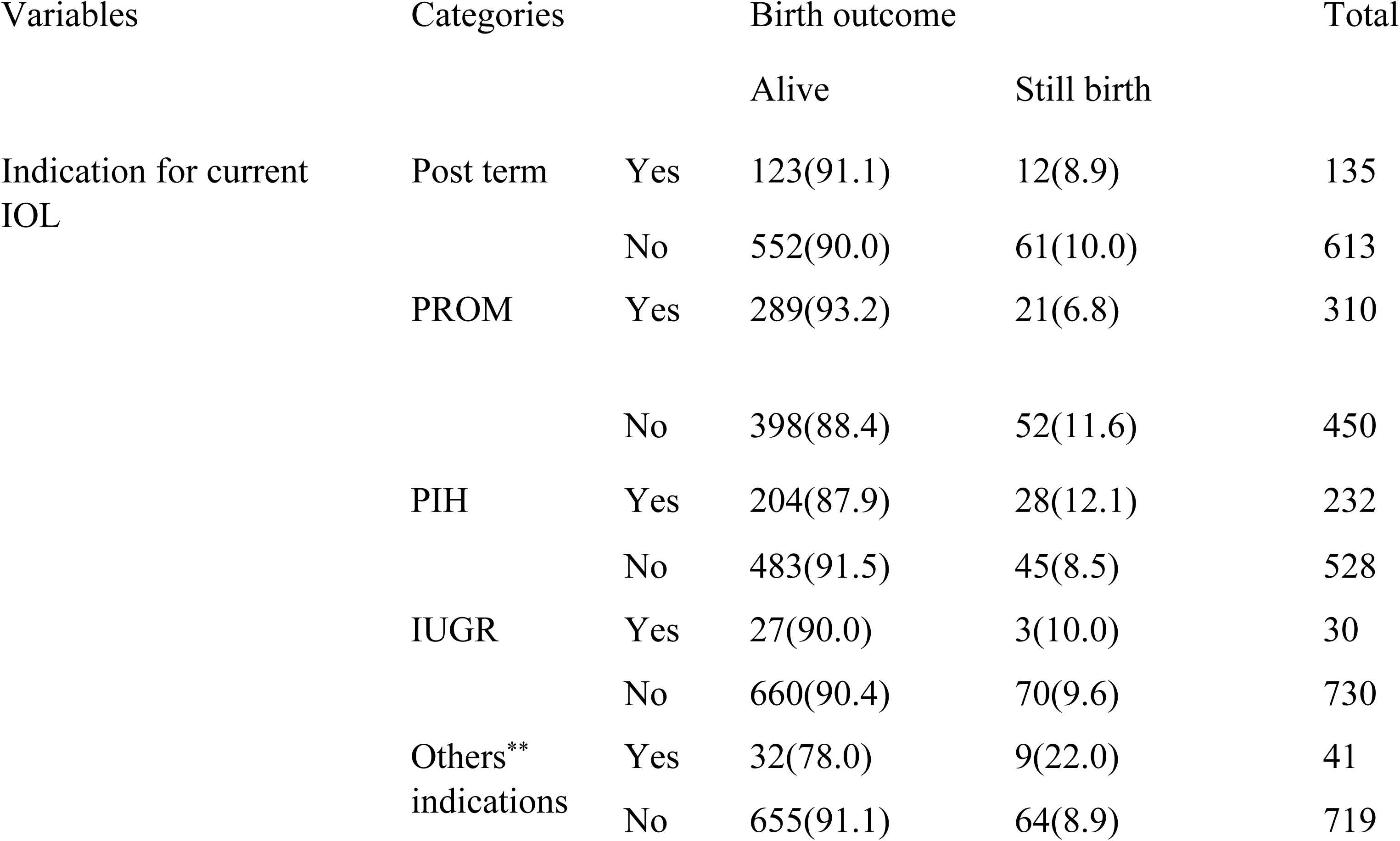
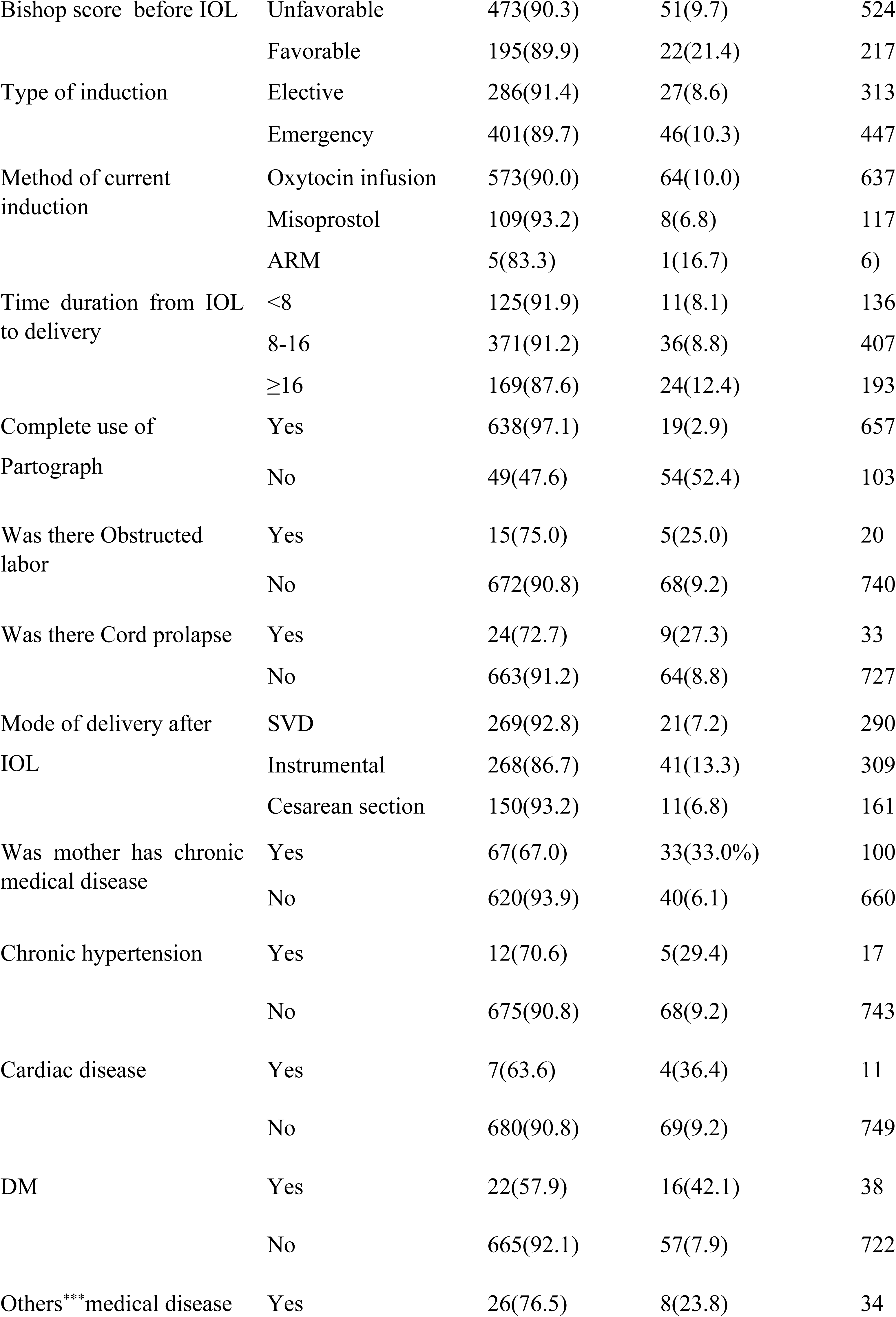
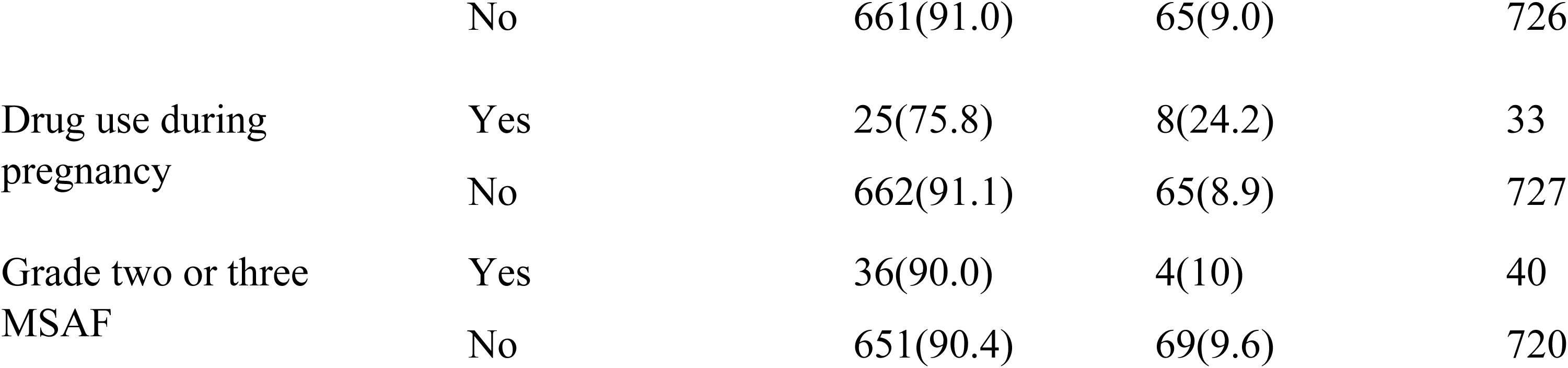
Induction and labor and delivery characteristics of women and new born after induction of labor in NEMMCSH south Ethiopia from January 01/2019 to December 31/2020 (n=760)

| Variables | Categories |  | Birth outcome |  | Total |
| --- | --- | --- | --- | --- | --- |
|  |  |  | Alive | Still birth |  |
| Indication for current IOL | Post term | Yes | 123(91.1) | 12(8.9) | 135 |
|  |  | No | 552(90.0) | 61(10.0) | 613 |
|  | PROM | Yes | 289(93.2) | 21(6.8) | 310 |
|  |  | No | 398(88.4) | 52(11.6) | 450 |
|  | PIH | Yes | 204(87.9) | 28(12.1) | 232 |
|  |  | No | 483(91.5) | 45(8.5) | 528 |
|  | IUGR | Yes | 27(90.0) | 3(10.0) | 30 |
|  |  | No | 660(90.4) | 70(9.6) | 730 |
|  | Others** indications | Yes | 32(78.0) | 9(22.0) | 41 |
|  |  | No | 655(91.1) | 64(8.9) | 719 |

|  |  |  |  |  |
| --- | --- | --- | --- | --- |
| Bishop score before IOL | Unfavorable | 473(90.3) | 51(9.7) | 524 |
|  | Favorable | 195(89.9) | 22(21.4) | 217 |
| Type of induction | Elective | 286(91.4) | 27(8.6) | 313 |
|  | Emergency | 401(89.7) | 46(10.3) | 447 |
| Method of current induction | Oxytocin infusion | 573(90.0) | 64(10.0) | 637 |
|  | Misoprostol | 109(93.2) | 8(6.8) | 117 |
|  | ARM | 5(83.3) | 1(16.7) | 6) |
| Time duration from IOL to delivery | <8 | 125(91.9) | 11(8.1) | 136 |
|  | 8-16 | 371(91.2) | 36(8.8) | 407 |
|  | ≥16 | 169(87.6) | 24(12.4) | 193 |
| Complete use of Partograph | Yes | 638(97.1) | 19(2.9) | 657 |
|  | No | 49(47.6) | 54(52.4) | 103 |
| Was there Obstructed labor | Yes | 15(75.0) | 5(25.0) | 20 |
|  | No | 672(90.8) | 68(9.2) | 740 |
| Was there Cord prolapse | Yes | 24(72.7) | 9(27.3) | 33 |
|  | No | 663(91.2) | 64(8.8) | 727 |
| Mode of delivery after IOL | SVD | 269(92.8) | 21(7.2) | 290 |
|  | Instrumental | 268(86.7) | 41(13.3) | 309 |
|  | Cesarean section | 150(93.2) | 11(6.8) | 161 |
| Was mother has chronic medical disease | Yes | 67(67.0) | 33(33.0%) | 100 |
|  | No | 620(93.9) | 40(6.1) | 660 |
| Chronic hypertension | Yes | 12(70.6) | 5(29.4) | 17 |
|  | No | 675(90.8) | 68(9.2) | 743 |
| Cardiac disease | Yes | 7(63.6) | 4(36.4) | 11 |
|  | No | 680(90.8) | 69(9.2) | 749 |
| DM | Yes | 22(57.9) | 16(42.1) | 38 |
|  | No | 665(92.1) | 57(7.9) | 722 |
| Others***medical disease | Yes | 26(76.5) | 8(23.8) | 34 |
|  | No | 661(91.0) | 65(9.0) | 726 |
| Drug use during pregnancy | Yes | 25(75.8) | 8(24.2) | 33 |
|  | No | 662(91.1) | 65(8.9) | 727 |
| Grade two or three MSAF | Yes | 36(90.0) | 4(10) | 40 |
|  | No | 651(90.4) | 69(9.6) | 720 |

### Neonatal characteristics after induction of labor

From the total induced pregnancies included in this study 15(2%) of the fetus developed non reassuring fetal heart rate pattern following induction. The mean (±SD) weight of the new born after induction of labor were 2755.97 grams ±614. In terms of sex of the new born 422(55.5%) were male. In other way above three fourth of the neonate 627(82.9) were had Apgar score seven and above during the fifth minute of delivery.

**Table 3:** Neonatal outcomes after induction of labor in NEMMCSH south Ethiopia from January 01/2019 to December 31/2020.

| Variable | Categories | Frequency | Percent |
| --- | --- | --- | --- |
| Outcome of the neonate | Alive birth | 687 | 90.4 |
|  | Still birth | 73 | 9.6 |
| Sex of the neonate | Male | 422 | 55.5 |
|  | Female | 338 | 45.5 |
| Apgar score at first minute | < 7 | 580 | 76.7 |
| | $\geq 7$ | 176 | 23.3 |
|  | No information | 4 |  |
| Apgar score at five minute | < 7 | 129 | 17.1 |
| | $\geq 7$ | 627 | 82.9 |
|  | No information | 4 |  |
| NRFHRP after IOL | Yes | 15 | 2 |
|  | No | 745 | 98 |
| Weight of the new born | ≤2499 | 265 | 35.1 |
|  | 2500-3999 | 454 | 60.2 |
|  | ≥4000 | 35 | 4.6 |
|  | No information | 6 |  |
| Need of NICU admission | Yes | 165 | 21.7 |
|  | No | 595 | 78.3 |

### Magnitude of Birth outcome after induction of labor

From the total 760 mothers who undergo induction of labor included in this study 73(9.6%) with 95% confidence level of (7.6, 11.7) mothers were gave still birth.

### Factors associated to birth outcome after induction of labor

In multivariate analysis residence, maternal chronic medical disease, previous history of still birth, Partograph use, and induction to delivery time were significant predictors.

The odd of acquiring still birth among mothers who were rural dwellers were had 3.59 times higher than those mothers who were urban dwellers (AOR=3.59; 95%CI: (1.32, 9.80)). The odd of acquiring still birth among mothers who had previous history of still birth were 7.45 higher than those mothers who hadn’t have previous still birth (AOR=7.45; 95%CI: (2.45, 22.38)). The odd of acquiring still birth among mothers who had chronic medical disease 3.58 times more likely than those mothers who hadn’t medical diseases (AOR=3.58; 95% CL: (1.23, 10.41)).The odd of acquiring still birth among mothers who gave birth less than eight hours and between eight and sixteen hours were 87% (AOR=0.13; 95%CI: (0.03,0.56)) and 72% (AOR=0.28; 95%CI:(0.10, 0.76)) less likely than those mothers who gave birth after sixteen hours respectively. The odd of acquiring still birth among mothers who were had complete Partograph follow up during active stage of labor were 96.6% less likely than their counterparts (AOR=0.034; 95%CI: 0.01,0.09).

**Table 4:**
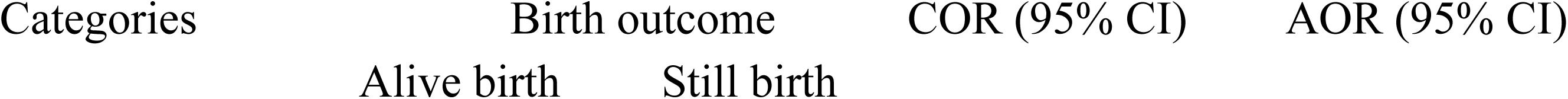
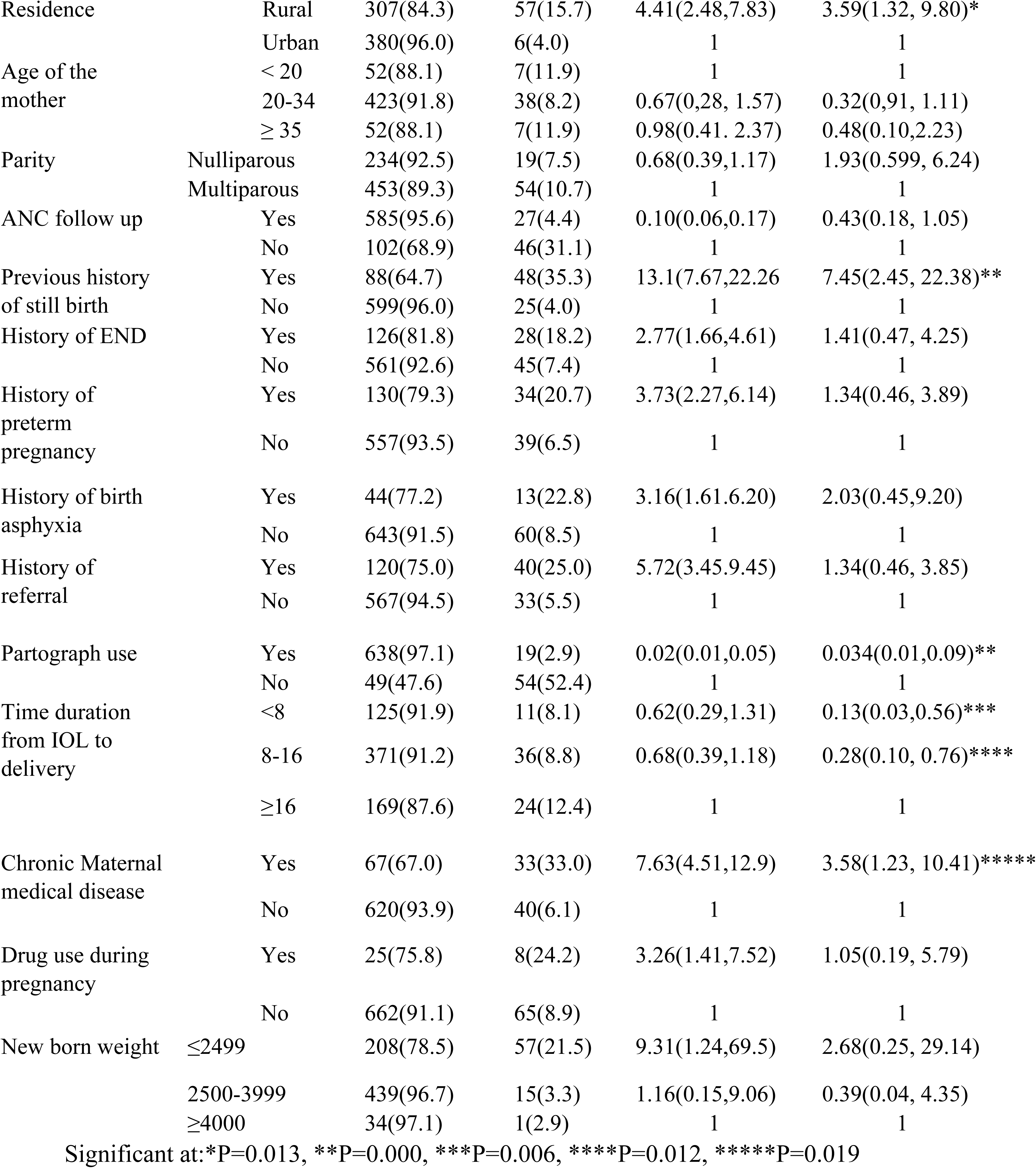
Factors associated with birth outcome after induction of labor in NEMMCSH south Ethiopia from January 01/2019 to December 31/2020 (n=760)

| Categories | Birth outcome |  | COR (95% CI) | AOR (95% CI) |
| --- | --- | --- | --- | --- |
|  | Alive birth | Still birth |  |  |

|  |  |  |  |  |  |
| --- | --- | --- | --- | --- | --- |
| Residence | Rural | 307(84.3) | 57(15.7) | 4.41(2.48,7.83) | 3.59(1.32, 9.80)* |
|  | Urban | 380(96.0) | 6(4.0) | 1 | 1 |
| Age of the mother | < 20 | 52(88.1) | 7(11.9) | 1 | 1 |
|  | 20-34 | 423(91.8) | 38(8.2) | 0.67(0.28, 1.57) | 0.32(0.91, 1.11) |
|  | ≥ 35 | 52(88.1) | 7(11.9) | 0.98(0.41, 2.37) | 0.48(0.10,2.23) |
| Parity | Nulliparous | 234(92.5) | 19(7.5) | 0.68(0.39,1.17) | 1.93(0.599, 6.24) |
|  | Multiparous | 453(89.3) | 54(10.7) | 1 | 1 |
| ANC follow up | Yes | 585(95.6) | 27(4.4) | 0.10(0.06,0.17) | 0.43(0.18, 1.05) |
|  | No | 102(68.9) | 46(31.1) | 1 | 1 |
| Previous history of still birth | Yes | 88(64.7) | 48(35.3) | 13.1(7.67,22.26) | 7.45(2.45, 22.38)** |
|  | No | 599(96.0) | 25(4.0) | 1 | 1 |
| History of END | Yes | 126(81.8) | 28(18.2) | 2.77(1.66,4.61) | 1.41(0.47, 4.25) |
|  | No | 561(92.6) | 45(7.4) | 1 | 1 |
| History of preterm pregnancy | Yes | 130(79.3) | 34(20.7) | 3.73(2.27,6.14) | 1.34(0.46, 3.89) |
|  | No | 557(93.5) | 39(6.5) | 1 | 1 |
| History of birth asphyxia | Yes | 44(77.2) | 13(22.8) | 3.16(1.61,6.20) | 2.03(0.45,9.20) |
|  | No | 643(91.5) | 60(8.5) | 1 | 1 |
| History of referral | Yes | 120(75.0) | 40(25.0) | 5.72(3.45,9.45) | 1.34(0.46, 3.85) |
|  | No | 567(94.5) | 33(5.5) | 1 | 1 |
| Partograph use | Yes | 638(97.1) | 19(2.9) | 0.02(0.01,0.05) | 0.034(0.01,0.09)** |
|  | No | 49(47.6) | 54(52.4) | 1 | 1 |
| Time duration from IOL to delivery | <8 | 125(91.9) | 11(8.1) | 0.62(0.29,1.31) | 0.13(0.03,0.56)*** |
|  | 8-16 | 371(91.2) | 36(8.8) | 0.68(0.39,1.18) | 0.28(0.10, 0.76)**** |
|  | ≥16 | 169(87.6) | 24(12.4) | 1 | 1 |
| Chronic Maternal medical disease | Yes | 67(67.0) | 33(33.0) | 7.63(4.51,12.9) | 3.58(1.23, 10.41)***** |
|  | No | 620(93.9) | 40(6.1) | 1 | 1 |
| Drug use during pregnancy | Yes | 25(75.8) | 8(24.2) | 3.26(1.41,7.52) | 1.05(0.19, 5.79) |
|  | No | 662(91.1) | 65(8.9) | 1 | 1 |
| New born weight | ≤2499 | 208(78.5) | 57(21.5) | 9.31(1.24,69.5) | 2.68(0.25, 29.14) |
|  | 2500-3999 | 439(96.7) | 15(3.3) | 1.16(0.15,9.06) | 0.39(0.04, 4.35) |
|  | ≥4000 | 34(97.1) | 1(2.9) | 1 | 1 |

Significant at: \*P=0.013, \*\*P=0.000, \*\*\*P=0.006, \*\*\*\*P=0.012, \*\*\*\*\*P=0.019

## Discussion

In this study the magnitude of still birth after induction of labor was 9.6% (95% CL: 7.6, 11.7). This result is greater than study done in Wolaita Sodo university referral hospital which was 6.1%, Aksum referral hospital (2.4%) and study done in Sudan (2.5%), (5, 15, 35) but the result of this is less than study done in Dessie referral hospital which is 21%, (24). The discrepancy for this result could be the sample size difference, socio demographic condition (70).

In this study the three predominant indications for induction were PROM 310(40.8%), PIH 232(30.5%) and post term 135 (17.8%) the result is in line with study done in India, Nepal, Jimma university specialized hospital, Wolliso St. Luke, Catholic Hospital and study done in Hawassa public hospitals (1,28,32,52),However study done in Wolaita Sodo University specialized hospital the three predominant indication for induction were PROM, post term and medical disorder of pregnancy while study done in Addis Ababa shows the predominant indication for induction were preeclampsia, PROM and post term pregnancy(15,27). The finding of this study also shows that the prevalence of failed induction was 22.1% which is in line with study done in Jimma university specialized hospital which was 21.4%(28) but this result was less than study done in Wolaita Sodo university teaching referral hospital which is 41.3% and study done Addis Ababa public hospital(15,27), The difference for this result could be due to failure in maintaining serum oxytocin concentration during the change of infusion bag or dose increment interval being 20 minutes which is not in accordance with currently accepted oxytocin pharmacokinetics (30).

In this study the two predominant methods of induction were oxytocin infusion 637(83.8%) and misoprostol 117(15.4%). This finding is supported by study done in Northern Tanzania which was from 1088 mothers who underwent IOL 82.08% of them were induced by oxytocin infusion and 13% were induced by prostaglandin, study done in Hawassa public hospitals and study done in Tigray Mekelle(31,40,55).

In this study the leading cause for neonatal intensive care unit admission was respiratory distress. This finding is supported by study done in Wolaita Sodo University teaching referral hospital in which neonatal distress were the leading cause for NICU admission (15).

The finding of this study also showed that the odd of getting still birth after induction of labor was 3.59 times [AOR=3.59; 95%CI: (1.32, 9.80)] more likely in rural dwellers mothers compared to urban dwellers. Even if this study include only induced mothers the finding is supported by study done in Amhara region (37). The reason for this might be mothers with rural residence have poor access to health information and there is also lake of transportation system to seek medical care and ANC follow up during pregnancy (21, 43).

The odd of getting still birth after induction of labor in this study was 7.45 times [AOR=7.45; 95%CI: (2.45, 22.38)] more likely in mothers who had previous history of still birth compared to those who hadn’t. The finding of this study is supported by study done in Nigeria which shows mothers with previous still birth were had more likely to have still birth[AOR= 3.79; 95%CL : (1.60–9.00)] (48). The reason for this might be related to maternal Rh-factor, which leads to erythroblastosis fatalis (71).

In this study the odd of acquiring still birth after labor induction on mothers who had chronic medical diseases were 3.58 times more likely [AOR=3.58; 95% CL: (1.23, 10.41)] compared to those mothers who hadn’t chronic medical diseases. This study is supported by study done in Tercha general hospital [AOR = 2.15; 95%CI :(1.28–3.62] (56). This might be maternal chronic and repeated pregnancy related comorbidities that result in fetal death (70).

Partograph utilization by service providers was the other strong factor associated with still birth. In this study mothers who were followed by Partograph during active phase of labor were 96.6% (AOR=0.034, 95%CL: 0.01, 0.09) less likely of getting still birth after induction of labor compared to their counterparts. The result is supported by study done in Aksum referral hospital which shows mothers who do not monitored by Partograph during labor were more likely to have still birth [AOR=8.66;95%CL: (2.88–26.10)] (16). This is due to poor Partograph utilization may resulted in depriving intervention like; cesarean section, instrumental delivery and early detection of maternal and neonatal complications or this might be due to use of Partograph can help alert health care providers to pick any abnormalities during the course of labor. Therefore, it can prevent perinatal loss with early diagnosis and management of labor complications (63).

In this study mother who were gave birth less than eight hours and between eight and sixteen hours were 87% (AOR=0.13; 95%CI: (0.03,0.56)) and 72% (AOR=0.28; 95%CI:(0.10, 0.76)) less likely getting still birth after labor induction respectively compared to those mothers who were gave birth above sixteen hours. This finding is supported by cross-sectional study done in Bonga General and Mizan Tepi University teaching hospitals which was showed mother who had labor length greater than 24 hours were 5 times more likely at risk of still birth than those who had labor length less than 24 hour (69). This might be due to prolonged labor increased risk of birth Aspasia, birth trauma, umbilical cord prolapse, PROM; which results increased perinatal mortality and morbidity (65).

## Conclusion

The magnitude of still birth after induction of labor was relatively high in the study area. Variables which increase the likelihood of still birth were, living in rural area, previous history of still birth and mothers with chronic medical diseases. However use of Partograph for assessing labor progress and giving birth before 16 hours after induction were decrease the likelihood of still birth. The determinants for still birth identified in this study can be prevented by providing appropriate care during antepartum and intrapartum period.

## Data Availability

All relevant data are within the manuscript and its Supporting Information files.

## Acknowledgments

The authors would like to acknowledge Wachemo University for financial assistance and Haramaya University for permission to conduct the thesis. Finally, authors would like to thanks study participants, data collectors and all concerned bodies who support us to conduct this thesis.

## Author contributions

DW, TA, TG, TK, were involved in the conception and design of the study, acquisition of data, analysis and interpretation of data, drafting and revising the article, agreed to submit to the current journal, gave final approval of the version to be published and agree to be accountable for all aspects of the work.

## Declarations

### Ethics approval and consent to participate

Ethical clearance was obtained from the Institutional Health Research Ethics Review Committee (IHRERC) Haramaya University, College of Health and Medical Sciences with Ethics approval number of IHRERC/074/2021. In addition, support letter was written to the hospital. The objective and purpose of the study were verified briefly to the study participant and confidentiality was assured. Finally, written informed consent was obtained from study participants before conducting the interview. This study was conducted in accordance with the Declaration of Helsinki.

### Consent for publication

Not applicable.

---

**Figure 1:**
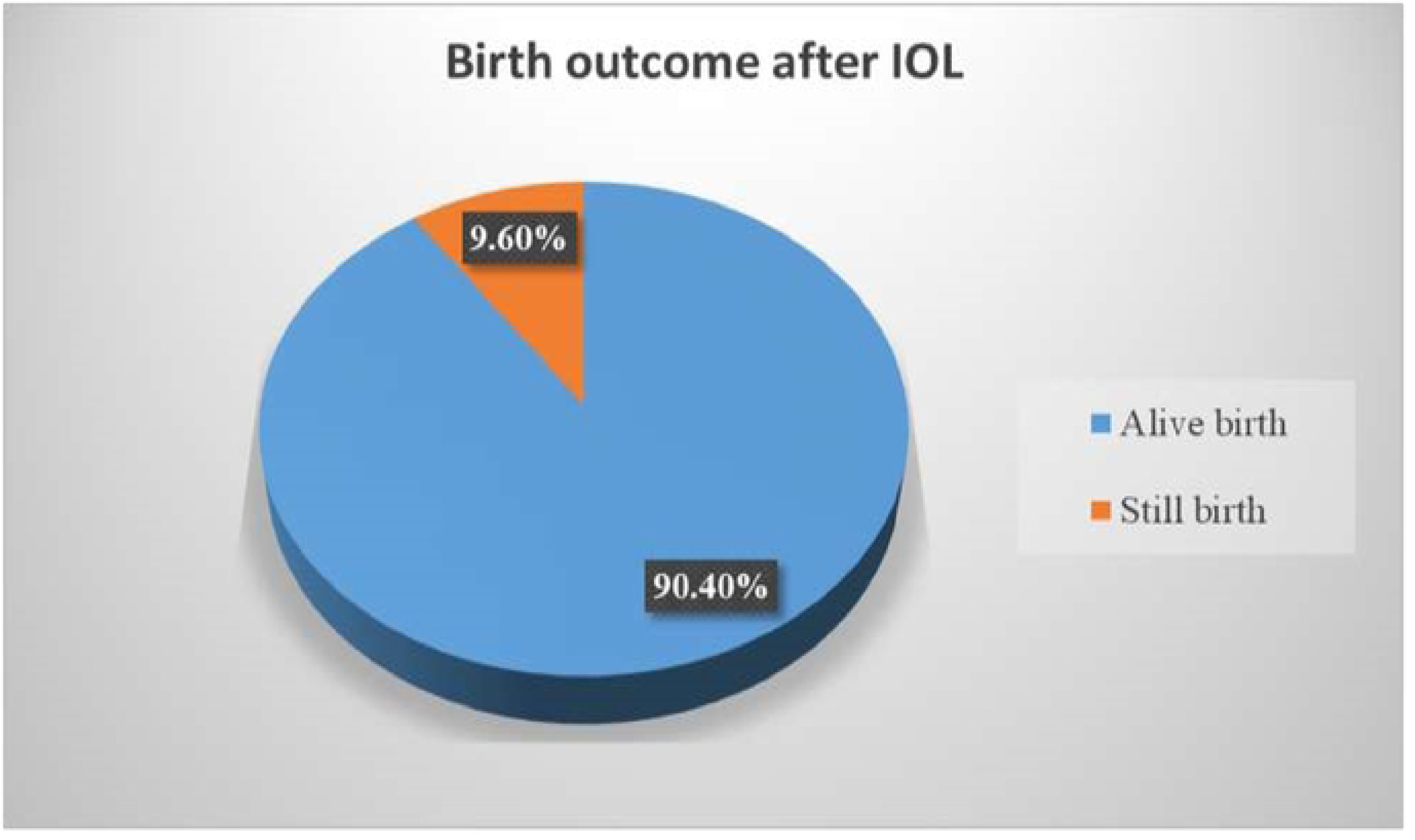
Birth outcome after induction of labor in NEMMCSH south Ethiopia from January 01/2019 to December 31/2020.

